# Developing implementation strategies to improve patient education for hypertension in primary care

**DOI:** 10.64898/2026.01.12.25341587

**Authors:** Kaylee Slater, Eleanor Clapham, Kim Beesley, Elizabeth Halcomb, Ben Kostyrka, Lisa Kouladjian O’Donnell, Liliana Laranjo, Florence Lopez, Mitchell Sarkies, Gautam Satheesh, Mouna Sawan, Catherine Stephen, John Stevens, Ritu Trivedi, Aletta E Schutte, Niamh Chapman

## Abstract

**Background:** Effective education improves hypertension self-management but is not routinely delivered, and there is limited guidance to overcome barriers to support delivery in primary care. This study aimed to use co-design and an implementation science framework to develop and prioritize implementation strategies for the delivery of patient education to support blood pressure (BP) management across the hypertension journey in primary care.

**Methods:** A two-stage, co-design study using implementation science frameworks. Stage 1 involved mapping interviews with patients to the Consolidated Framework for Implementation Research and Expert Recommendations for Implementing Change to generate strategies to improve patient education for BP management along the care journey (diagnosis, treatment initiation, long-term management). Stage 2 involved a three-hour multi-disciplinary workshop to refine, rank and prioritize these strategies using patient personas, clinical scenarios, and participatory voting. Finally, the strategies were reviewed by consumer advisors with lived experience of BP management to ensure suitability and acceptability.

**Results:** Six context-specific strategy prompts aligned to stages of the hypertension journey were paired with patient personas and clinical scenarios to guide workshop discussions. Twenty-one participants (n=2 general practitioners, n=4 nurses, n=6 pharmacists, n=10 industry professionals) attended the workshop. Ultimately, six refined strategies included: (1) in-pharmacy education for suitable device purchase using a structured protocol; (2) general practitioner consultations about home BP measurement with nurse follow-up; (3) nurse-led drop-in group education sessions; (4) nurse-led group education on interpreting readings with general practitioner follow-up; (5) nurse-led lifestyle planning with allied health referrals; and (6) pharmacist-delivered BP measurement and medication review. Pharmacy-based strategies (1) and (6) ranked highest for feasibility and impact, and cost-effectiveness, respectively.

**Conclusion:** We identified evidence-based multidisciplinary strategies to improve the delivery of patient education for hypertension, co-designed with clinicians. Future work should focus on evaluating their implementation in real-world primary care settings.

**KEY MESSAGES:** *What is already known on this topic:* Patient education improves hypertension self-management, yet it is rarely delivered consistently in primary care and there is limited guidance on how to overcome practical barriers to implementing education strategies across the care journey.

*What this study adds:* This study co-designed and prioritized six multidisciplinary, context-specific strategies to deliver patient education for blood pressure management at diagnosis, treatment initiation, and long-term care, using implementation science frameworks.

*How this study might affect research, practice or policy:* These strategies offer a practical roadmap for embedding structured education into routine primary care by leveraging existing workforce and infrastructure, aligning with funded models, and supported by clinician incentives to enhance uptake.

## 1 INTRODUCTION

Hypertension is the leading modifiable risk factor for death and disability, affecting 1.2 billion people worldwide (1). Globally, only 21% of individuals with hypertension achieve blood pressure (BP) control (1) despite affordable and well-tolerated therapies (2). Effective BP management requires accurate measurement, sustained lifestyle risk reduction and lifelong adherence to prescribed antihypertensive medications (3, 4). This is best achieved via multi-disciplinary care and effective patient education to support self-management (4–6). Patient education plays a critical role in encouraging active engagement in care by providing individuals with the knowledge needed to make informed decisions about their health and adopt effective self-management behaviours, such as lifestyle changes and home BP monitoring (7). Patients who receive education for hypertension management have greater self-efficacy, adherence to medication and uptake of lifestyle changes, to support hypertension management (8).

Effective patient education for hypertension utilises a multifaceted approach that combines personalized content for patient needs with tailored and integrated feedback, and can be delivered in various formats such as print, digital, and face-to-face interactions (9, 10). Implementing effective patient education for hypertension requires a comprehensive approach that begins with a thorough assessment of patient needs (11). This involves understanding preferences regarding content, delivery methods, cultural backgrounds, and health literacy levels (11) to ensure that the education is accessible to all patients throughout all stages of the hypertension care journey.

Hypertension is the most commonly managed condition in primary care (12) and patients value receiving health information directly from health professionals (13, 14). Interventions to improve BP control are more effective when they adopt a multidisciplinary team-based care approach that actively involves doctors as well as pharmacists and nurses (8). However, current models of care often place limited emphasis on patient education, and busy health practitioners are often challenged to incorporate education into their practice within resource constraints. Therefore, it is important to understand how standardised delivery of patient education for hypertension can be effectively integrated into primary care settings.

To address the growing burden of hypertension and increasing workforce pressures in primary care, there is an urgent need to integrate multidisciplinary team-based care models that make better use of existing workforces and provide structured, consistent education across the hypertension care journey (15). Therefore, this study aimed to use an implementation science framework and co-design to develop and prioritise implementation strategies for the delivery of patient education to support BP management across the hypertension journey in primary care.

## 2 METHODS

### 2.1 Study design

A two-stage, sequential co-design study using implementation science frameworks was conducted to generate, refine and prioritize implementation strategies (Figure 1). Stage 1 involved mapping interviews with patients to implementation science frameworks to generate the strategies, while Stage 2 involved a multi-disciplinary workshop with primary care practitioners and industry representatives to refine, rank and prioritize these strategies. The implementation strategies were iteratively reviewed by consumer advisors with lived experience of hypertension management to ensure suitability and acceptability. Human Research Ethics Committee of the University of Tasmania (H0028867) and the Human Research Ethics Committee of the University of Sydney (2024/HE000528) gave ethical approval for this work. This study was reported as per the Standards for Reporting Implementation Studies Statement (StaRI) (16).

**Figure 1.**
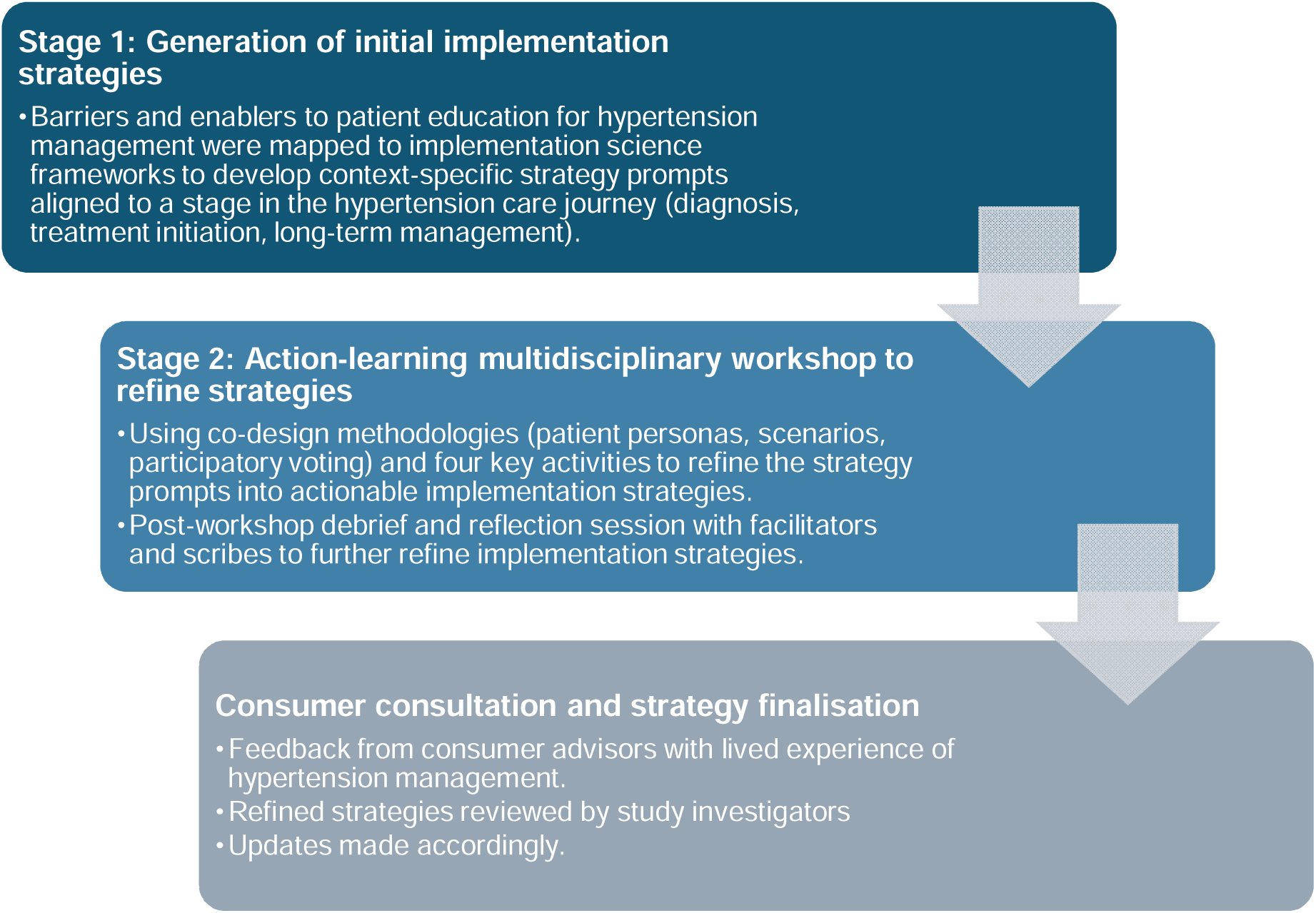
Study design

### 2.2 Stage 1: Generation of initial implementation strategies

Semi-structured interviews with patients were conducted as part of a previous study to explore barriers and enablers to patient education for BP management across the hypertension care journey. The detailed methodology, including participant eligibility, recruitment, data collection and analysis, is published elsewhere (17). In brief, interviews with adults ≥18 years who had measured BP at home in the past 12 months were analysed using Framework Analysis to identify determinants of patient education delivery across three stages of care (diagnosis, treatment initiation, and long-term management) (17). These findings, along with patient personas and clinical scenarios developed in that study, informed the initial implementation strategies for this work.

Barriers and enablers identified through this analysis were deductively mapped to the Theoretical Domains Framework (18) (Supplementary File 1) and to the 2022 Consolidated Framework for Implementation Research (CFIR) (19) to identify relevant constructs across individual, organisational, and system levels (Supplementary File 2). CFIR constructs were then manually mapped to relevant Expert Recommendations for Implementing Change (ERIC) strategies using the most up-to-date CFIR-ERIC mapping tool to produce an output of the most to least expert-endorsed *Level 1* and *Level 2* strategies (20). The ERIC framework includes 73 discrete strategies designed to address known implementation barriers (20). In this study, strategies with a cumulative mapping frequency >75% were selected and synthesised into actionable and context-specific strategy prompts to take forward to Stage 2. One investigator completed data mapping (KS), meeting regularly with other study investigators (NC, EC) to discuss findings and resolve any concerns.

### 2.3 Stage 2. Multidisciplinary workshop to refine and prioritize strategies

#### Participants and recruitment

A purposive sampling approach was used to recruit primary care professionals (pharmacists, doctors (general practitioners), and general practice nurses) and health industry representatives. Participants were recruited either through existing networks, Facebook Advertisement or publicly available staff directories. To be included, healthcare practitioners needed to be currently practising in Australia, and providing care related to BP management in a primary care setting. Industry representatives were required to be employed in organisations with a relevant focus (such as pharmacy advocacy, public health services, non-government health organisations) and interest in improving patient education for BP management. Informed consent was gained via an e-consent process. Health professionals were reimbursed with an electronic gift card for their attendance.

#### Data collection

The three-hour in-person workshop was held in July 2025 at The University of Sydney, Australia. Guided by an Action-Learning framework (21) and co-design principles, the workshop used experiential, problem-solving methods to enable participants to identify barriers, enablers, and solutions related to strategy implementation. Workshop activities included structured group exercises for strategy refinement, plenary feedback discussions, and participatory voting using hearts, stars and dollar signs to assess the perceived feasibility, impact, and cost-effectiveness, respectively, of each strategy (see Supplementary File 3 for details). Participants were grouped by profession with industry representatives distributed across the groups. Each group was assigned one patient persona and clinical scenario with an accompanying strategy prompt, supported by a facilitator (a clinician-researcher from the same profession) and a scribe who documented discussions using structured templates. At the workshop conclusion, a debrief and reflection session was held with all facilitators and scribes to support researcher reflexivity and capture observations on group dynamics, engagement, and emerging insights.

#### Data analysis

Qualitative data from table discussions documented by the scribes and the workshop audio recordings were analysed using a thematic content analysis approach. Similar suggestions were combined and collapsed into a single suggestion and a summary of all unique suggestions per strategy were combined into a final strategy summary. As recommended by previous research to report implementation strategies, final strategies were reported in six dimensions: actor, the action, action targets, temporality, dose, and implementation outcomes addressed (22).

Quantitative data were analysed using descriptive statistics (n, %) for each strategy where the higher number of stars and hearts indicated better feasibility and impact, respectively, and the lower number of dollar signs indicated better return-on-investment. To rank the strategies and account for missing data in the number of responses per strategy, weighted average scores were calculated using number of votes for each rating (e.g. how many people gave each score) to calculate the average to ensure that scores reflected the full distribution of responses, not just the most common or total number. Min-max normalisation was then applied to compare feasibility, impact, and cost on the same scale. A composite score for each summary was calculated using the formula *composite score = normalised feasibility + normalised impact – normalised cost* to identify strategies that were rated as highly feasible and impactful while also being cost-effective. Lastly, ideas were ranked based on their composite scores to identify the most to least preferred strategies.

#### Consumer consultations

Consumer advisors (n=4) met virtually for one-hour before the workshop (May 2025) and for two-hours after the workshop (August 2025) to inform revisions to language, hypertension care journey and perceived feasibility of proposed and revised strategies. The first meeting aimed to familiarise consumers with study methods and outline of the workshop activities. Ahead of the second meeting, refined implementation strategies were sent via email for independent review to ensure consumer feedback was appropriately applied. The second meeting gathered structured feedback on the relevance and acceptability of the refined strategies using facilitated discussion by KS to determine if the if key time-points in hypertension care were captured and if the refined strategies addressed barriers experienced in accessing patient education for hypertension. This feedback was incorporated into the final versions, and the final six strategies were reviewed and approved by all members of the authorship team. Both meetings were recorded, and field notes were taken by study investigators (KS, NC, EC).

## 3 RESULTS

### 3.1 Stage 1

Ten relevant CFIR constructs were identified and manually mapped to ERIC implementation strategies. We identified 43 Level 1 and Level 2 ERIC strategies (Supplementary File 4), of which 16 ERIC strategies were retained for further development. Of these, 10 ERIC strategies were synthesised into six actionable and context-specific strategy prompts (Supplementary File 5). Each strategy prompt addressed one or more barriers to delivering standardised BP education and was aligned to a specific stage in the hypertension journey (diagnosis, treatment initiation, or long-term management). Each strategy was paired with a patient persona and clinical scenario to support contextualisation and facilitate discussion in the workshop (Supplementary File 6). Figure 2 summarises the six strategy prompts, including the stage of the hypertension care journey it targets. The remaining six ERIC strategies informed the design of the Stage 2 workshop and consumer engagement processes.

**Figure 2.**
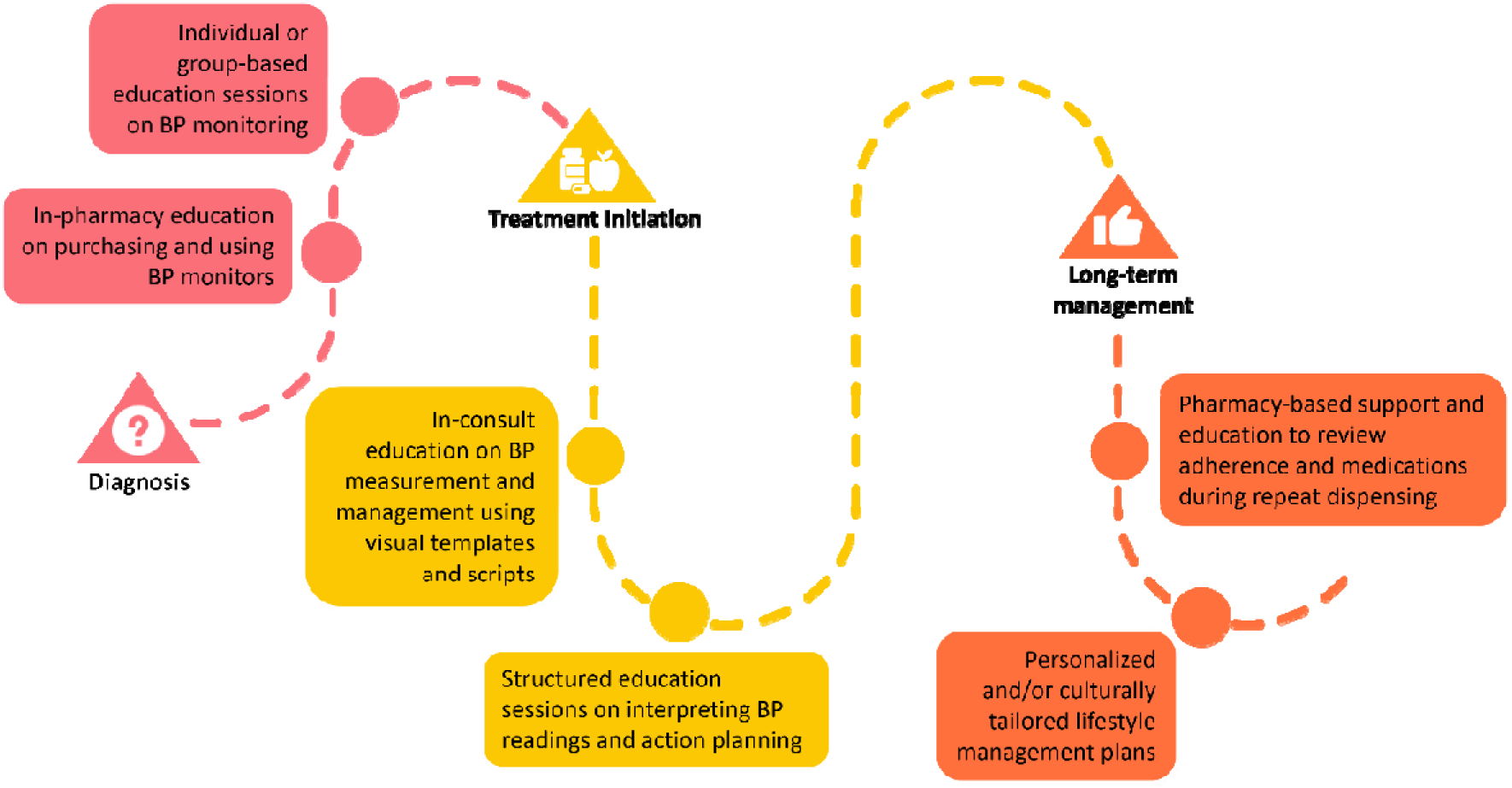
Summary of the strategy prompts taken forward to the workshop. BP: blood pressure.

### 3.2 Stage 2

#### 3.2.1 Participants

Twenty-one participants attended the workshop, of whom 11 (52%) were practicing health professionals and 10 (48%) were industry representatives (Supplementary File 7). Most were female (n=16, 76%), based in New South Wales (n=19, 90%), and more than half (n=12, 57%) had less than 5 years of experience in their current role. Industry representatives worked at not-for-profit research or industry advocacy organisations, pharmaceutical and medical device companies, primary health organisations and local health districts.

#### 3.2.2 Challenges to implementation of the strategies

Participants brainstormed a diverse range of challenges to implementing the six strategy prompts for BP education in primary care (Table 1). Time and workload pressures were consistently cited across all strategy prompts, with practitioner participants highlighting limited consultation duration and competing demands as constraints on delivering quality education. Funding and resourcing limitations were also raised, particularly around remuneration for nurse and pharmacist-led education, patient access to validated home BP monitors for self-monitoring, and culturally appropriate materials. Low perceived patient motivation and health literacy were viewed as challenges to engaging patients, often tied to the asymptomatic nature of hypertension. Coordination gaps between pharmacies and general practices were also seen as key challenges to continuity of care.

**Table 1.** Challenges to implementation of strategies to improve BP patient education.

| Challenge | Description |
| --- | --- |
| Time and workload pressures | Limited consultation time, competing priorities, and busy practice environments impede the ability to deliver patient education. |
| Funding constraints | Inadequate funding for nurse or pharmacist-led services. |
| Patient motivation and engagement | Patients show low ownership, resistance to education, and poor understanding due to the asymptomatic nature of hypertension. |
| Educational resource limitations | Lack of accessible, appropriate, and culturally sensitive materials. |
| Role and scope clarity | Ambiguity around how pharmacists and nurses can contribute and underutilization of skills. |
| Coordination and communication gaps | Limited information sharing and poor coordination between doctors, nurses, and pharmacists undermines continuity of care. |
| Biases and professional assumptions | Health professionals may assume patients are already educated or adherent or show limited motivation to provide education. |
| Technology and infrastructure gaps | Limited interoperability, lack of structured data capture tools, and absence of telehealth options in pharmacies. |
| Cultural and language barriers | Inadequate language support and lack of culturally sensitive approaches to education. |
| Professional trust and motivation | Some nurses and pharmacists feel undervalued or not trusted to deliver education. |

BP: blood pressure.

#### 3.2.3 Refining implementation strategies

In response to the identified challenges, participants collaboratively refined their strategy prompts into detailed delivery plans and feedback discussions revealed recurring concerns related to funding, feasibility, and integration into clinical practice. Ultimately, through further refinement, six strategies were produced, each tailored to address the specific challenges identified and designed to embed structured education within existing workflows, leverage team-based care, and enhance patient engagement. Table 2 summarises the strategies and the challenges they aim to overcome. Of the six strategies, two focused on improving patient uptake and use of validated home BP monitors, including pharmacist-delivered education at the point of device purchase using a structured protocol, and a dedicated doctor’s consult supported by digital educational materials to educate patients on correct measurement technique and enable telemonitoring. Nurse-delivered strategies included group drop-in sessions and one-on-one consultations to provide education, combining motivational interviewing, action-planning, and provision of educational materials tailored to patients’ literacy and cultural needs. Group education sessions delivered online, or in-person focused on helping patients interpret home BP readings, with follow-up doctor consultations and decision support embedded within digital health tools. Two strategies also addressed long-term BP management, including nurse-delivered personalised and/or culturally appropriate lifestyle plans, and pharmacist-delivered BP medication reviews conducted during repeat dispensing to review medication adherence and reinforce BP education. Collectively, these strategies emphasised personalized, team-based care, integration with existing systems, and the use of digital tools to support ongoing self-monitoring and engagement.

**Table 2.** Education implementation strategies for improving the delivery of patient education for BP management.

| Strategy | Actor/s | Action/s | Target/s of the action | Temporality | Dose | Barriers addressed |
| --- | --- | --- | --- | --- | --- | --- |
| **BP monitor education protocol offering a 4-step structured protocol ("I Heart 1–4") at pharmacies** | Pharmacist and pharmacy assistants | • Needs assessment: understand what the device will be used for.<br>• Budget alignment: match devices to patient affordability while ensuring quality.<br>• Device validation: teach patients how to identify clinically validated machines.<br>• Hands-on demonstration: show patients how to operate their monitor correctly, supported by simple information sheets. | Patients purchasing and using home BP monitors | At time of purchase | Single session (with optional follow-up) | Time constraints in consultations, resource limitations and professional assumptions |
| **Doctor consult with the BP Toolkit for BP measurement education** | Doctors and practice nurses | • Doctor educates patients on measurement technique and accurate recording using BP Toolkit digital tools and provides validated device list.<br>• Patients learn to log their home BP readings into the BP Toolkit.<br>• A follow-up appointment with a practice nurse is offered 1-week later to review technique and readings. | Patients newly diagnosed with hypertension | At diagnosis and 1-week follow-up | Two sessions (initial with doctor and optional follow-up with nurse) | Limited consultation time; health professional coordination gaps; lack of standardised resources |
| **Face-to-face education package combining group drop-in sessions and individual support.** | Nurses | • Structured group “drop-in” face-to-face BP measurement education sessions led by nurses after referral from the patient’s doctor.<br>• Patients receive home BP kits which include guidance on validated low-cost BP monitors,<br>printed or digital resources from BP Toolkit detailing practical instruction for undertaking and reporting home measurement. | Patients at diagnosis or with low health literacy needing additional support | At diagnosis or as needed (drop-in model) | One group session (with optional follow-up) | Low health literacy; patient disengagement; resource and material limitations |
| **Nurse delivered, doctor-advocated group education session on interpreting home BP readings and action-planning.** | Nurses (delivery) and doctors (referral) | • Group (hybrid-offering) education session to teach patients about interpreting their home-BP readings.<br>• Patients will receive access to the BP Toolkit to help troubleshoot their home BP readings and make informed decisions about their care.<br>• Patients to also attend a follow-up doctor consult within 3-months of the group session. | Patients needing support interpreting home BP | Early after diagnosis with doctor follow-up within 3 months | One session and ongoing Toolkit access, 1 doctor follow-up | Low health literacy; limited patient coordination between health professionals |
| **Nurse-delivered lifestyle plan and allied health referral** | Nurses | • Tailored BP management education with a printed care-plan for the patient.<br>• Referral to allied health for more lifestyle intervention with a chronic disease management plan (small out-of-pocket cost).<br>• Direct patient to the BP Toolkit for further resources. | Patients requiring lifestyle change and/or culturally tailored plans | As part of chronic disease management plan | Single session, linked to care planning cycle and referral development | Cultural/language barriers including limited appropriate resources; fragmented information |
| **Pharmacist-delivered BP medication review during dispensing** | Pharmacist and pharmacy assistants | • BP measurement done by a trained pharmacy assistant<br>• Medication review by the pharmacist to assess medication adherence and provide BP management education. | Patients on $\geq 1$ BP medication | During repeat dispensing | 20-45 minutes offered annually | Underutilisation of pharmacists, poor continuity of care, time constraints in practice |

BP: blood pressure.

#### 3.2.4 Implementation strategy ranking and prioritisation

Participants consistently emphasised feasibility and cost-effectiveness when refining strategies and favoured those that leveraged existing funding structures such as nurse-delivered chronic disease management plans and pharmacist-led medication reviews. Group-based education was viewed as a cost-effective use of clinician time, reducing patient costs while enhancing peer support and engagement. Incentives for both clinicians (e.g., remuneration) and patients (e.g., low or no-cost options) were seen as critical for uptake and sustained delivery. Strategies that improved coordination among doctors, nurses, and pharmacists rather than introducing entirely new programs were perceived as more sustainable. These perspectives informed the ranking outcomes, with pharmacy-based strategies scoring highest for feasibility, impact, and return on investment.

Table 3 displays the normalised results of strategy ranking for feasibility, impact and return on investment from most preferred (first) to least preferred (sixth). One strategy received the best score for both feasibility and impact (pharmacist-delivered BP medication review), and a different strategy received the best score for return-on-investment (four-step BP monitor education protocol at pharmacies). The least preferred strategy for feasibility and impact was a nurse-delivered lifestyle plan with an accompanying allied health referral, and for return-on-investment was a dedicated doctor consultation for home BP measurement counselling.

**Table 3.** Comparative ranking of strategies by multidisciplinary workshop attendees.

| Strategy | Ranking |  |  |  |
| --- | --- | --- | --- | --- |
|  | Overall | Feasibility | Impact | Return on investment |
| Pharmacist-delivered BP medication review during dispensing | **1** | 1 | 1 | 3 |
| Face-to-face education package combining group drop-in sessions and individual support. | **2** | 2 | 2 | 4 |
| Four-step BP monitor education protocol at pharmacies | **3** | 4 | 5 | 1 |
| Doctor consult with the BP Toolkit for BP measurement education | **4** | 3 | 4 | 6 |
| Nurse delivered, doctor-advocated group education session on interpreting home BP readings and action-planning. | **5** | 5 | 3 | 5 |
| Nurse-delivered lifestyle plan and allied health referral | **6** | 6 | 6 | 2 |

BP: blood pressure. Strategies are listed in descending order of preference, where 1 represents the highest-ranked strategy and 6 the lowest.

## 4 DISCUSSION

This study employed rigorous implementation science and co-design methods to refine and prioritize strategies for improving patient education in hypertension management. Grounded in the CFIR-ERIC frameworks, the approach integrated patient perspectives through patient personas and clinical scenarios and stakeholder perspectives from doctors, nurses, pharmacists, and industry representatives in an action-learning workshop. This structured, participatory process enabled the identification of practical, context-specific solutions and the development of six tailored strategies that embed patient education into routine care, spanning diagnosis to long-term management, and aligned with both patient needs and the operational realities of primary care.

Growing evidence supports shifting patient education delivery beyond medically focused models to include nurse and pharmacist-led approaches for hypertension (23, 24). Time constraints of doctors in primary care represents a considerable obstacle to patient education, which is the cornerstone of BP management to ensure medication adherence and self-management (23, 24). When refining the strategies, participants suggested that pharmacist and nurses’ skills are underutilized in BP management, and therefore five of the six strategies were proposed to be delivered by either pharmacists or general practice nurses. Community pharmacies are an ideal setting to support a multidisciplinary approach to improve BP management and one that is cost-effective for health services and patients (25, 26), although are currently disconnected from general practice. Pharmacist-led approaches, including medication counselling, education, and support for home BP monitoring have demonstrated reductions in BP and improved medication adherence (23). Similarly, nurse-led interventions have shown benefits for BP control, lifestyle modification, and medication adherence (27). While participants supported nurse and pharmacist-led care, they emphasised the critical role of doctors in initiating and coordinating referrals to practice nurses. Evidence suggests that hypertension management is most effective when delivered through team-based models of care, with coordinated contributions from diverse health professionals (23). This highlights the need for coordinated team-based education supported by shared resources along the entire hypertension care journey.

Delivery of patient education for hypertension management is constrained by several systemic and structural barriers, such as limited time and training to adapt education to patients’ health literacy needs, and by a lack of coordinated, multimodal communication to support ongoing learning and follow-up (28–30). To address these, our strategies emphasise structured, role-specific follow-up and multimodal education, combining verbal delivery with accessible resources like printed or digital education materials. A multimodal approach to delivering patient education reinforces key messages delivered during consultations thereby extending the reach of education beyond the clinical encounter (9). Providing tangible resources alongside active verbal delivery of patient education also addresses variations in health literacy, language, and learning preferences, making patient education more effective (31).

Incorporating culturally tailored education and digital tools into revised care strategies helps address longstanding gaps in culturally responsive care, where standard education often fails to meet the diverse cultural, linguistic, and lifestyle needs essential for effective behaviour change (32). Participants and consumers strongly advocated for group-based education and lifestyle support to be embedded throughout the care journey. These group-based interventions foster peer support and shared learning, which have been shown to improve BP outcomes (33) and are particularly effective in culturally and linguistically diverse populations (34). However, this workshop did not specifically address implementation strategies for culturally and linguistically diverse populations and dedicated engagement with diverse populations and context-specific exploration will be required in future work.

### Strengths and limitations

This study employed a robust co-design approach that incorporated diverse professional and consumer perspectives, and utilised persona-based methods grounded in implementation science frameworks. Extensive preparatory work ensured that strategy prompts were evidence-informed even prior to the workshop. Nonetheless, the findings of this study should be interpreted considering several limitations. While the implementation strategies were developed using evidence-based frameworks, co-design methodology, and qualitative data, there was a small and uneven sample across professions at the workshop. Pharmacists were over represented, while doctors, and health professionals with more than 20-years’ experience were under-represented. These factors may have introduced a level of bias and limit generalizability to routine care. Additionally, this study was conducted within the Australian context, with discussions shaped by local health system structures and funding constraints. Future research should consider expanding stakeholder engagement to include more experienced doctors, nurses and international and diverse perspectives. Further consumer workshops could also help assess the acceptability and perceived impact of proposed strategies with diverse consumers, including prioritization based on patient preferences.

### Implication considerations and future directions

Our implementation strategies have strong potential for integration into primary care by leveraging the existing workforce and infrastructure, aligning with currently funded models, and supported by clinician incentives to enhance uptake. However, policy action is needed to enable and sustain team-based education models, particularly nurse and pharmacist-led approaches underpinned by interprofessional collaboration and integration of digital tools. Embedding tailored and structured education across the care journey is essential to ensure effective hypertension management. Beyond their immediate application, the methods used to develop and refine the strategies also provide a replicable, context-sensitive approach for co-designing implementation strategies in primary care, offering valuable contributions to implementation science and chronic disease management.

## Conclusions

This study used implementation science frameworks and co-design principles to develop, refine and prioritize implementation strategies to improve the delivery of patient education for hypertension in primary care. The refined strategies emphasised role-specific interventions embedded in existing workflows in general practice and pharmacies, focusing on home BP monitoring, the use of digital tools, group education sessions and tailored education to support personalized, team-based care for BP management. These findings contribute to the growing body of evidence supporting the implementation of patient education in primary care settings. Grounded in input from community members, clinicians, and experts, the proposed strategies offer actionable, evidence-based guidance for practice. Moreover, they provide a foundation for enhancing patient education across a range of chronic conditions and comorbidities, reinforcing the value of integrated, context-sensitive approaches in primary care.

## Sources of funding

This work was supported by multiple funding sources; National Health and Medical Research Council of Australia Investigator Grant (GNT2018077), Foundation for High Blood Pressure Research Early Career Research Transition Grant 2023-2024 and New South Wales Health, Office for Health and Medical Research (DG23/7050). MS is funded by an NHMRC Investigator Grant (2007970) and Sydney Horizon Fellowship. LL is funded by a National Health and Medical Research Council Investigator Grant (2017642) and Sydney Horizon Fellowship. AS is funded by an Investigator Grant from the National Health and Medical Research Council of Australia (APP2017504).

## Competing interests

No relevant disclosures.

## Data availability

The data that support the findings of this study are available from the corresponding author upon reasonable request.

## Author contributions

Conceptualisation, KS, EC and NC.; methodology, KS, EC and NC; formal analysis, KS, EC and FL; writing – original draft preparation, KS, EC; writing – review and editing, KS, EC, CB, EH, BK, LKO, LL, FL, MS, GS, MS, CS, RT, AES and NC; supervision, NC; project administration, KS. All authors have read and agreed to the final version of the manuscript.

## Acknowledgements

We would like to sincerely thank our consumer advisors, John Stevens, Kim Beesley and William Steele, as well as one advisor who preferred to remain anonymous for their invaluable insights, feedback, and contributions to this work. Their perspectives greatly informed the development, refinement and applicability of the implementation strategies.

## SUPPLEMENTARY FILES

**Supplementary File 1.** Barriers to the delivery of patient education for blood pressure measurement and management

**Supplementary File 2.** Mapping identified barriers to CFIR constructs

**Supplementary File 3.** Multidisciplinary Workshop Activities

**Supplementary File 4.** Level 1 and 2 Strategies Identified by updated CFIR-ERIC Manual Matching Tool

**Supplementary File 5.** Overall strategy summaries mapped to ERIC strategies, patient hypertension journey and delivery mode

**Supplementary File 6.** Summary of the strategy prompts aligned to patient personas which were taken forward to the workshop

**Supplementary File 7.** Demographic characteristics of multidisciplinary workshop attendees

**Supplementary File 8.** Plain Language Summary

## Notes

### Competing Interest Statement

The authors have declared no competing interest.

### Author Declarations

Human Research Ethics Committee of the University of Tasmania (H0028867) and the Human Research Ethics Committee of the University of Sydney (2024/HE000528) gave ethical approval for this work.

