## Supplementary Files for "Developing implementation strategies to improve patient education for hypertension in primary care"

**Supplementary File 1.** Barriers to the delivery of patient education for blood pressure measurement and management

**Supplementary File 2.** Mapping identified barriers to CFIR constructs

**Supplementary File 3.** Multidisciplinary Workshop Activities

**Supplementary File 4.** Level 1 and 2 Strategies Identified by updated CFIR-ERIC Manual Matching Tool

**Supplementary File 5.** Overall strategy summaries mapped to ERIC strategies, patient hypertension journey and delivery mode

**Supplementary File 6.** Summary of the strategy prompts aligned to patient personas which were taken forward to the workshop

**Supplementary File 7.** Demographic characteristics of multidisciplinary workshop attendees

**Supplementary File 8.** Plain Language Summary

**Supplementary File 1.** Barriers to the delivery of patient education for blood pressure measurement and management

| Barrier | Summary explanation | TDF | Exemplar quotes |
| --- | --- | --- | --- |
| 1 | Pharmacists often lack training, resources, and education necessary to provide detailed guidance on selecting blood pressure monitors and educating patients on measurement. Additionally, there is a common assumption that patients already know how to measure their blood pressure correctly, resulting in minimal instructions being given, which may compromise effective self-monitoring. | Knowledge  Skills  Professional Role and Identity  Beliefs about capabilities  Environmental Context and Resources | *" I know Omron is good, but these days there's just so many different ones. It really depends on their budget, like how, how much they can afford basically." [Pharmacist, female, practising 5-9y]*  *"Home measurement probably is a lesser part of it. I would say. I probably provide less education about that. Unless a patient has asked or prompted more about it." [GP, female, practising 5-9y]*  *“They said yeah, “this one is good buy that one. And I think they might have said, just put it around, press the button”. I think that's about all we got told at the chemist.” [Patient, male, 50-59y]* |
| 2 | Patients receive inconsistent and primarily verbal information about home blood pressure measurement during consultations, with full instructions rarely provided. The absence of high-quality, standardised templates and resources for recording, interpreting, and learning measurement techniques further limits patients’ ability to manage their blood pressure effectively. | Knowledge  Skills  Environmental Context and Resources  Memory, Attention and Decision Processes | *"But if that person had, like a blood pressure monitor that they have access to, then I would just get them to do the worksheet within that week as well and then check their blood pressure again." [GP, female, practising 5-9y]*  *“My doctor did show me or give me instructions about how to do it and make sure I knew about how to best arm down, which my sister-in-law did as well. [Patient, female, 50-59y]*  *“OK, taking the reading, he gave me a printed hardcopy spreadsheet that from memory allowed to me to record it. Maybe three recordings morning and three recordings in the afternoon. Something like that for seven days and basically said good luck and bring this back.” [Patient, male, 60-69y]*  *"Normally, a doctor recommends that [home BP monitoring] to patients. Yeah, patients have, like issue with their blood pressure. They-patients normally have the monitoring device at home, and they'll just say “Oh, yeah, but we're at home like, I monitor it regularly”. Well, we don't normally-doctors said-like they generally wouldn't say that or give you that information." [Nurse, female, practising <5y]*  *"He just said I'll just do it a couple of times a day if you want, it was pretty vague. So I sort of just decided to do it and it in my way.” [Patient, female, 40-49y]* |
| 3 | Patients rely on observing healthcare professionals or seeking online resources to learn how to measure their blood pressure, which may lead to inconsistent or incomplete training and affect the accuracy and confidence of self-monitoring. The lack of standardised instruction from HCPs can hinder effective self-management. | Skills  Knowledge  Behavioural Regulation (habits)  Environmental Context and Resources  Belief about Capabilities | *"But I think most of the people that do it, I think, have a good idea of how to do it. Because they are putting it on their arm… I don't know whether they're putting on their left or their right arm, but they're generally putting it above their elbow crease." [Pharmacist, female, practising 16-20y]*  *“Yeah, I do it the way I've seen them do it. I use the way you see professionals doing it, so I imitate that.” [Patient, female, 40-49y]*  *"We wouldn't…they wouldn't know how to fit on the cuffs, perhaps, so they could forget. And you never know what they're doing at home if they're measuring It's a bit tricky to say, for sure what's going on back at home. [Nurse, female, practising <5y]* |
| 4 | Lack of patient education on blood pressure thresholds and uncertainty about appropriate actions following high readings. This results in patients feeling that their concerns are dismissed and that home blood pressure monitoring is intended primarily for healthcare professionals rather than as a tool for patients' long-term management. | Knowledge  Skills  Beliefs about Capabilities  Beliefs about Consequences  Professional role and identity. | *"We do notice, a lot of patients do take the blood pressure before they come in, and they always let us know that “Oh we just want to compare with your machine”. And it seems to be accurate so far. And so I just go based off what they've said as well." [Nurse, female, practising <5y]*  *"I find in the older age group, either that they have easier access to those things already. They're better able to do like multiple readings over a few days, and sometimes that can be actually that can add a little bit more information as well." [GP, female, practising 5-9y]*  *“From what I've read is the 140/90... Once you sort of start going over those measurements, you're starting to get into maybe see your GP about it. Umm and yeah, normally when I've gone because I’m getting, you know, let's call it the 140 / 90 or higher, I'll go and see them.” [Patient, male 70-79y]* |
| 5 | Lack of standardised information provided to patients, leading many to seek out information on their own. This self-directed approach can result in inconsistent understanding, although patients express a preference for trustworthy and reputable sources, such as credible websites. The absence of structured guidance from healthcare professionals may hinder patients' confidence and ability to manage their condition effectively. | Knowledge  Environmental Context and Resources  Memory, Attention and Decision Processes  Beliefs about capabilities. | *“I go to Stroke Foundation now. Stroke Foundation has, has a pretty big library, and it's a good resource that you get there. So look at the stories that we look at situations where somebody like me has been facing a similar situation.” [Patient, male, 40-49y]*  *"Yes, yeah, definitely. Yeah, I think you're not going to achieve patient change unless they have that health literacy that's present there and know what things that they need to change, and you can't just do it with medication alone. And I think the patient has a better experience when they've got more education." [GP, female, practising 5-9y]*  *"If English is their first language, that makes it a lot easier. If they are linguistically diverse, it's culturally and linguistically diverse, it's obviously a lot harder... But sometimes it's very hard to find things within that with realm. I find it very hard to get things." [GP, female, practising 10-15y]*  *“Even still today I get a bit confused, and I get varied responses, and it depends on whom I'm talking to now. I still really don't know what a good normal reading is.” [Patient, male, 40-49y]* |
| 6 | Limited time and competing priorities faced by GPs however limited utilisation of other primary health practitioners (nurses, pharmacists). These constraints reduce opportunities for regular BP measurement and for delivering structured patient education, which can negatively impact patients' understanding and engagement in managing their condition. | Environmental Context and Resources  Professional Role and Identity | *"Generally speaking, people are pretty good with their blood pressure medication. But obviously, if they're on like multiple different medications for different other health conditions. It can be a little bit overwhelming when there's lots of changes or additions to a medication regime. So that's when we would probably flag that sort of thing." [Pharmacist, female, practising 16-20y]*  *"Oh, It's tricky. I don't think they really notify us whether or not they took it from the pharmacy. I haven't encountered any instance of such." [Nurse, female, practising <5y]*  *"Yeah. Oh, I mean, it would probably be good if we had a system where we could have like more blood pressure check-ins outside of just the doctor's appointment." [GP, female, practising 5-9y]*  *“I can't imagine that they'd like you doing that (bringing your BP readings to the doctor) given that they don't actually have much time.” [Patient, female, 60-69y]*  *“And I think some of the benefits of home BP are that it's way more accurate than the GP doing it. And the reason is that the GPs blood pressure monitor looks like it came out of the ark. And they always take my blood pressure and over my clothes and they always talk to me and asked me to speak back to them. They're asking because they're in short, you know, they're time poor.” [Patient, female 60-69y]* |

BP: blood pressure. HCPs: healthcare professionals. GP: general practitioner. TDF: Theoretical Domains Framework

**Supplementary File 2.** Mapping identified barriers to CFIR constructs

| **Barrier elements** | **Barrier** | **CFIR Domain** | **CFIR Construct** | **Explanation** |
| --- | --- | --- | --- | --- |
| Pharmacists lack training/resources to provide BP monitor education | 1 | Inner setting | Available resources | Lack of dedicated resources like training, educational materials, and staff time to support proper implementation. |
|  |  |  | Access to Knowledge and Information | Pharmacists don’t have ready access to digestible, accurate resources to support BP monitor education. |
| Health professionals assume patients know how to measure BP and give minimal instructions | 1-3 | Inner setting | Culture: recipient-centredness | Assuming patients can accurately measure BP without guidance reflects a low patient-centred culture, as it overlooks their actual needs and may compromise the effectiveness of self-monitoring and management. |
| Limited standardisation and completeness in delivery of patient education by HCPs. | 2-4 | Inner setting | Structural characteristics | Organisational setup (e.g., clinic workflow) limits GPs’ ability to spend time or provide detailed BP education. |
|  |  |  | Access to knowledge and Information | GPs don’t have easy access to high-quality educational resources/templates to use in-consultations. |
|  |  |  | Communications | Poor communication structures between HCPs and patients may lead to inconsistency in patient education messages |
| Inconsistent tools to support patient understanding and self-management of blood pressure | 2, 5 | Inner setting | Available resources | Staffing limitations (e.g., lack of time or funding) to spend time educating patients and a lack of materials to use educational tools limits the ability to provide thorough standardised education. |
| Patients’ ability to measure and manage BP is limited by unstandardised patient education | 1-4 | Individuals (patients) | Capability | Patients need to have the knowledge and skills for proper BP monitoring. |
|  |  |  | Opportunity | Patients need supportive environmental conditions (e.g., recording templates and educational resources). |
|  |  |  | Motivation | Patients must be motivated to engage in BP monitoring/management, which can be influenced by the quality of education and support received |
|  |  | Implementation | Innovation recipients | Patients require standardised and good quality patient education in order to manage BP. |
| Patients lack the knowledge about BP thresholds and how to action high BP readings at home | 4 | Individuals (patients) | Capability | Inadequate patient education results in patients lacking the knowledge and skills to interpret BP readings and decide next steps without their HCPs. Reduces patient engagement and activation. |
|  |  | Inner setting | Access to knowledge and information | Patients and providers lack clear, accessible educational resources explaining BP targets and follow-up actions. |
| HCPs are prioritising in-clinic BP measurements over home BP monitoring, leaving patients feeling that their home readings are being dismissed. | 4 | Inner setting | Communications | The quality and nature of communication between patients and providers related to their home BP readings is leaving patients feeling unheard. |
|  |  |  | Relative priority | Home BP monitoring is not seen as important compared to in-clinic measures, reducing support and education for home BP monitoring. |
|  |  | Individuals (patients) | Motivation | Feeling dismissed reduces patients’ motivation to engage with self-monitoring and management. |
|  |  |  | Opportunity | Lack of external support that frames home BP monitoring as a patient-empowering behaviour reduces patients perceived opportunity to engage. |
| Patients resort to self-directed information seeking, risking inconsistent or inaccurate knowledge | 5 | Individuals (patients) | Capability | Patients vary in their ability to discern credible information and incorporate it correctly. |
|  |  |  | Opportunity | A lack of guidance and delivery of credible information and resources to patients reduces the opportunity to use the information, especially for those who cannot access it independently. |
|  |  |  | Motivation | Patients’ preference for credible sources demonstrates motivation, however, the absence of guidance may lower their confidence and self-efficacy. |
| Limited GP time and competing priorities hinder the delivery of standardised and complete patient education for BP management | 6 | Inner setting | Available resources | GPs face constraints on time, staffing, and other resources, limiting their capacity to perform regular BP measurement and deliver comprehensive patient education. |
|  |  |  | Relative priority | Within primary care, BP education and home monitoring may not be viewed as highly important relative to other urgent clinical tasks, reducing focus and effort. |
|  |  |  | Structural characteristics | The organization’s workflows, staffing models, and task distribution patterns may not support delegation or structured delivery of BP education, constraining how services are provided. Nurses are not being utilised for education delivery. |
| Pharmacists and nurses are not effectively engaged in delivering BP education in primary care | 6 | Inner setting | Relational connections | Lack of effective team coordination between GPs, and nurses restricts the shared delivery of patient education. |
|  |  |  | Communications | Lack of effective communication channels and document sharing or referral processes between GPs, pharmacists, and nurses also restricts the shared delivery of patient education. |
|  |  |  | Culture | The prevailing norms and assumptions within general practice does not prioritise delegation of education tasks to non-GP providers, limiting multidisciplinary approaches. |
|  |  | Outer setting | Partnerships and connections | The extent of relationships between GP practices and external organisations like pharmacies are hindering collaborative BP education and shared management of patients. |

BP: blood pressure. GP: general practitioner

### **Supplementary File 3.** Multidisciplinary Workshop Activities

*Activity 1: Strategy refinement.* Participants reviewed their assigned persona and scenario and using sticky notes identified challenges to implementation. Using think-aloud discussion, they then collaboratively identified solutions to implementation by detailing how it could be delivered (HOW), by whom (WHO), what resources were required (WHAT), when it could realistically be implemented (WHEN), why this strategy is important in improving patient education (WHY) and estimated cost (HOW MUCH).

*Activity 2: Group feedback.* One member from each group had three minutes to present their refined strategy to the full group. A seven-minute facilitated plenary discussion followed, allowing cross-table feedback and idea exchange. Scribes recorded feedback for integration into subsequent refinements.

*Activity 3: Final refinement and elevator pitch.* Groups reconvened to incorporate feedback provided by the group discussion and further refine their strategy. Using think-aloud discussion, participants detailed what successful implementation would look like by considering feasibility, impact and cost-effectiveness. The scribe detailed a summary of the refined strategy and one member from each group did a 90-second elevator pitch to the full group.

*Activity 4: Participatory voting.* In an interactive exercise, participants individually evaluated each strategy using a sticker-based voting system. Each participant ranked feasibility, impact and cost-effectiveness from 1(lowest)-6(highest). This was inverted for cost-effectiveness where six represented least cost-effectiveness (most expensive to implement) and one represented most cost-effective.

**Supplementary File 4.** Level 1 and 2 Strategies Identified by updated CFIR-ERIC Manual Matching Tool

| **ERIC strategies** | **Cumulative** | Available resources | Access to knowledge and information | Communications and Relational connections | Relative priority | Structural characteristics | Culture | Recipient-centredness | Partnerships and connections | Innovation recipients |
| --- | --- | --- | --- | --- | --- | --- | --- | --- | --- | --- |
| Capture and share knowledge | **147%** | 22% | 31% | 26% |  | 23% | 22% |  | 23% |  |
| Access for readiness and identify barriers and facilitators | **146%** |  |  |  | 36% | 36% | 41% | 33% |  |  |
| Create a learning collaborative | **141%** |  | 45% | 35% |  |  | 30% |  | 31% |  |
| Promote network weaving | **130%** |  |  | 57% |  | 23% |  |  | 50% |  |
| Involve patients and family members | **130%** |  |  |  |  |  |  | 71% |  | 59% |
| Build a coalition | **128%** |  |  | 39% |  | 27% |  |  | 62% |  |
| Identify and prepare champions | **126%** |  | 24% |  |  | 27% | 52% |  | 23% |  |
| Conduct local consensus discussions | **119%** |  |  | 22% | 46% |  | 22% | 29% |  |  |
| Use advisory boards and workgroups | **118%** |  |  |  |  |  | 22% | 29% | 35% | 32% |
| Obtain and use patients and family feedback | **117%** |  |  |  |  |  |  | 76% |  | 41% |
| Conduct local needs assessment | **111%** |  |  |  | 32% |  | 22% | 57% |  |  |
| Prepare patients to be active participants | **103%** |  |  |  |  |  |  | 48% |  | 55% |
| Conduct educational meetings | **101%** |  | 79% |  |  |  | 22% |  |  |  |
| Develop educational materials | **86%** |  | 59% |  |  |  |  |  | 27% |  |
| Change physical structure and equipment | **80%** | 48% |  |  |  | 32% |  |  |  |  |
| Access new funding | **78%** | 78% |  |  |  |  |  |  |  |  |
| Intervene with patients to enhance uptake and adherence | **74%** |  |  |  |  |  |  | 24% |  | 50% |
| Develop resource sharing agreements | **57%** | 26% |  |  |  |  |  |  | 31% |  |
| Facilitation | **56%** |  |  | 26% |  |  | 30% |  |  |  |
| Alter patient fees | **55%** | 22% |  |  |  |  |  |  |  | 23% |
| Distribute educational materials | **55%** |  | 55% |  |  |  |  |  |  |  |
| Organise clinician implementation team meetings | **52%** |  |  | 52% |  |  |  |  |  |  |
| Conduct educational outreach visits | **51%** |  | 28% |  |  |  |  |  | 23% |  |
| Develop academic partnerships | **50%** |  |  |  |  |  |  |  | 50% |  |
| Promote adaptability | **45%** |  |  |  |  | 23% | 22% |  |  |  |
| Inform local opinion leaders | **44%** |  |  | 22% |  |  | 22% |  |  |  |
| Use mass media | **41%** |  |  |  |  |  |  |  |  | 41% |
| Fund and contract for clinical innovation | **39%** | 39% |  |  |  |  |  |  |  |  |
| Alter incentive or allowance structure | **39%** |  |  |  | 39% |  |  |  |  |  |
| Conduct ongoing training | **38%** |  | 38% |  |  |  |  |  |  |  |
| Visit other sites | **38%** |  |  |  |  |  |  |  | 38% |  |
| Recruit, designate and train for leadership | **33%** |  |  |  |  |  | 33% |  |  |  |
| Mandate change | **32%** |  |  |  | 32% |  |  |  |  |  |
| Tailor strategies | **30%** |  |  |  |  |  | 30% |  |  |  |
| Increase demand | **29%** |  |  |  | 29% |  |  |  |  |  |
| Centralise technical assistance | **26%** |  |  | 26% |  |  |  |  |  |  |
| Provide local technical assistance | **24%** |  | 24% |  |  |  |  |  |  |  |
| Conduct cyclical small tests of change | **23%** |  |  |  |  | 23% |  |  |  |  |
| Involve executive boards | **23%** |  |  |  |  |  |  |  | 23% |  |
| Identify early adopters | **23%** |  |  |  |  | 23% |  |  |  |  |
| Make billing easier | **22%** | 22% |  |  |  |  |  |  |  |  |
| Use other payment schemes | **22%** | 22% |  |  |  |  |  |  |  |  |
| Shadow other experts | **21%** |  | 21% |  |  |  |  |  |  |  |

Level 1: strategies with majority (over 50%) endorsement and Level 2: Top quartile overall involved 20% or more endorsement.

The updated CFIR (2022) includes constructs that aligned closely with our data, including communications, relational connections, and COM-B-based individual patient determinants (capability, opportunity, and motivation). However, these newer constructs are not incorporated into the updated CFIR-ERIC matching tool. To inform the selection of relevant implementation strategies, we aligned communications and relational connections with networks and communications from the 2009 CFIR, based on conceptual overlap. The COM-B-based constructs were pragmatically mapped to related 2022 CFIR domains, access to knowledge and information, available resources, and recipient-centeredness, to reflect patients’ capacity and readiness to the delivery of patient education for BP management.

**Supplementary File 5.** Overall strategy summaries mapped to ERIC strategies, patient hypertension journey and delivery mode

|  | **Strategy Summary** | **Journey stage** | **Delivery mode** | **ERIC Strategies** | **Justification** |
| --- | --- | --- | --- | --- | --- |
| 1 | Pharmacists provide education on purchasing and using BP monitors | Diagnosis | Active | - Conduct educational meetings - Develop educational materials - Prepare patients to be active participants - Identify and prepare champions | Focus on training pharmacists, providing tools, and encouraging patient engagement. |
| 2 | Individual or group-based education sessions on BP monitoring led by trained nurses | Diagnosis | Active | - Conduct educational meetings - Identify and prepare champions - Prepare patients to be active participants - Create a learning collaborative - Access new funding | Formalises nurse involvement with structured sessions. |
| 3 | In-consult GP education on BP measurement and management using visual templates and scripts | Diagnosis/ Treatment Initiation | Active | - Conduct educational meetings - Develop educational materials - Prepare patients to be active participants - Identify and prepare champions | Standardises GP-led education at the start. |
|  | Personalised self-monitoring tools and action plans distributed in consults | Treatment Initiation | Passive | - Develop educational materials - Change physical structure and equipment - Prepare patients to be active participants | Passive reinforcement of messages via handouts. |
| 4 | GP-led structured education sessions on interpreting BP readings and action planning | Treatment Initiation | Active | - Conduct educational meetings - Prepare patients to be active participants - Involve patients and family members | Targets early improvements in health literacy for treatment and adherence. |
|  | Provision of printed and/or digital education tools to patients | Treatment Initiation | Passive | - Develop educational materials - Promote network weaving - Change physical structure and equipment | Leverages communication infrastructure for passive delivery. |
| 5 | Nurse-led personalised and/or culturally tailored lifestyle management plans | Treatment Initiation | Active | - Prepare patients to be active participants - Involve patients and family members - Conduct educational meetings - Create a learning collaborative | Empowers patients through tailored planning support. |
|  | Online patient education materials promoted and available via trusted public sources (Heart Foundation, clinic waiting rooms, social media) | Long-term Management | Passive | - Promote network weaving - Develop educational materials - Change physical structure and equipment - Build a coalition - Access new funding | Widespread dissemination and patient access beyond the clinic environment. |
| 6 | Pharmacy-based support and education to review adherence and medications during repeat dispensing | Long-term management | Active | - Conduct educational meetings - Build a coalition - Identify and prepare champions - Prepare patients to be active participants - Promote network weaving | Normalise and structure pharmacists’ role in long-term support. |

BP: blood pressure. GP: general practitioner

**Supplementary File 6.** Summary of the strategy prompts aligned to patient personas which were taken forward to the workshop

|  | Persona | Summary | Stage | Delivery | Strategy summary | Rationale | Barriers addressed |
| --- | --- | --- | --- | --- | --- | --- | --- |
| 1 | Margaret (F, 50-59y) | An accountant recently surprised by a high BP reading, feeling anxious and overwhelmed by home monitoring and choosing a device. | Diagnosis | Active | Pharmacists provide education on purchasing and using BP monitors | Margaret is overwhelmed by BP monitor choices and lacks device literacy. Pharmacist-led education directly supports her at the point of purchase. | Pharmacists lack home BP monitoring resources; assumptions that patients already know how to measure BP. |
| 2 | Bruce (M, 70-79y) | Limited health literacy, recently diagnosed but likely hypertensive for years, struggling with accurate BP monitoring and in need of basic, supportive education. | Diagnosis | Active | Individual or group-based education sessions on BP monitoring led by trained nurses | A structured session is crucial for reinforcing correct measurement and correcting misconceptions. | Inconsistent verbal-only education; lack of standardised templates and resources; patients rely on observation or online info. |
|  |  |  |  | Passive | Provision of printed and/or digital education tools to patients | Repeated, embedded education helps reinforce learning in future consults without overwhelming him |  |
| 3 | Nick (M, 60-69) | A stressed project manager with a family history of hypertension, motivated to monitor his BP and improve his health but lacks continuity of care from the same practitioner, and is uncertain about measurement technique hesitant about starting medication. | Treatment initiation | Active | In-consult GP education on BP measurement and management using visual templates and scripts | Structured education reassures Nick, offers clarity, and addresses his hesitation around medication with facts. | Uncertainty about BP thresholds and next steps; lack of standardised instruction instructions on home BP monitoring; lack of standardised templates and resources. |
|  |  |  |  | Passive | Personalised self-monitoring tools and action plans distributed in consults | A take-home resource gives Nick something to refer to outside appointments to consolidate his understanding. |  |
| 4 | Helen (F, 60-69y) | A retired educator who monitors her BP regularly but lacks understanding of what her readings mean and feels her home monitoring is overlooked by her GP. | Treatment initiation | Active | GP-led structured education sessions on interpreting BP readings and action planning | Addresses uncertainty about medications, fluctuating readings, and lack of explanation at visits. | Patients have uncertainty about BP thresholds and next steps; lack of feedback from health professionals. |
| 5 | Amina (F, 40-49y) | A community worker from a CALD background who takes medication consistently but struggles with dietary changes and fragmented information, seeking culturally relevant guidance. | Long-term management | Active | Nurse-led personalised and/or culturally tailored lifestyle management plans | Culturally tailored support helps Amina manage lifestyle-related challenges and stay engaged. | Patients seek information independently; lack of culturally relevant, structured guidance. |
|  |  |  |  | Passive | BP Toolkit promoted and available via trusted public sources (Heart Foundation, clinic waiting rooms, social media) | Amina actively seeks information online, however trusted, culturally inclusive public resources will help her find what she’s looking for. |  |
| 6 | Elias (M, 60-69y) | A retired engineer who manages his BP with medication but has become less active and forgetful over time, and would benefit from updated, evidence-based support for long-term management. | Long-term management | Active | Pharmacy-based support and education to review adherence and medications during repeat dispensing | Elias sees his pharmacist regularly; leveraging that touchpoint supports adherence and monitoring. | Limited time in GP consults; underutilisation of pharmacists and nurses for education. |

BP: blood pressure. CALD: culturally and linguistically diverse. GP: general practitioner

**Supplementary File 7.** Demographic characteristics of multidisciplinary workshop attendees

| **Demographic characteristics** | **n (%)** |
| --- | --- |
| Profession |  |
| Industry representative | 10 (48) |
| General practitioner/doctor | 2 (10) |
| Registered nurse | 4 (19) |
| Community pharmacist | 5 (24) |
| Gender | |
| Male | 5 (24) |
| Female | 16 (76) |
| Age | |
| 18-39 years | 12 (57) |
| 40-59 years | 9 (43) |
| Location | |
| New South Wales | 19 (90) |
| Victoria | 2 (10) |
| Years in practice/industry | |
| <5 years | 12 (57) |
| 5-9 years | 5 (24) |
| 10-15 years | 2 (10) |
| 16-20 years | 2 (10) |

**Supplementary File 8.** Plain Language Summary

Education helps people care for high blood pressure but is not often delivered well in primary care. There is also little guidance on how to overcome problems to delivering education well.

This study aimed to co-design strategies to improve how patient education is delivered across the hypertension primary care journey. This includes at diagnosis, when starting treatment, and long-term care.

**How this study was conducted**

We used a two-stage process.

Stage 1: Interviews with patients were mapped to implementation science frameworks to find barriers and create enabling strategies.

Stage 2: A three-hour workshop with 21 (general practitioners, nurses, pharmacists, and industry professionals) to refine and prioritise these strategies.

**Key Findings**

Six practical strategies were created to improve how education is given to patients in primary care:

1. In-pharmacy education on buying and using the right blood pressure device.
2. GP sessions supported by an online blood pressure education platform with nurse follow up.
3. Nurse-led drop-in group education sessions.
4. Nurse-led group sessions on how to read and action home blood pressure readings with follow up by a general practitioner.
5. Nurse-led lifestyle planning with allied health follow up.
6. Pharmacist-delivered blood pressure measurement and medication review.

**Take-home message**

We created six strategies to improve patient education for blood pressure management in primary care. The next step is to test these strategies in real-world settings.
